# Prediction and prognosis of delayed cerebral ischemia via continuous monitoring of blood-brain barrier permeability

**DOI:** 10.1101/2023.02.12.23285830

**Authors:** Chao Zhang, Wenjuan Tang, Liang Cheng, Chen Yang, Ting Wang, Juan Wang, Zhuang Miao, Xintong Zhao, Xinggen Fang, Yunfeng Zhou

**Affiliations:** Departments of Radiology, Yijishan Hospital of Wannan Medical College, Wuhu 241001, Anhui, China; Departments of Neurosurgery, Yijishan Hospital of Wannan Medical College, Wuhu 241001, Anhui, China

**Author notes:** Correspondence: Yunfeng Zhou, Department of Radiology, Yijishan Hospital of Wannan Medical College, No.2 Zheshan west Road, Wuhu, 241001, China. Chao Zhang and Wenjuan Tang contributed equally to this work.

## Abstract

**Background:** Blood-brain barrier disruption is a prominent pathological characteristic of aneurysmal subarachnoid hemorrhage (aSAH), which can be measured as K^trans^ using CT perfusion.

**Purpose:** To monitor K^trans^ within 24 hours of aSAH and during the time window associated with a delayed cerebral ischemia (DCI) event (DCITW), and to explore its association with the trajectory of DCI, including outcome at three months.

**Methods:** We retrospectively assessed consecutive aSAH patients from a prospective database between July 2020 and September 2022. Patients were grouped according to the DCI occurrence and three months modified Rankin scale. K^trans^ at admission (admission K^trans^) and during DCITW (DCITW K^trans^) were compared between DCI and non-DCI groups, and between good outcome and poor outcome groups. The changes in K^trans^ were also analyzed. Multivariate logistic regression analysis was performed to identify independent predictors of DCI and poor outcome.

**Results:** One hundred and twenty-eight patients (mean age, 61±12 [SD]; 75 women) were included. Both admission K^trans^ (0.58±0.18 vs 0.47±0.12, *P*=0.002) and DCITW K^trans^ (0.54±0.19 vs 0.41±0.14, *P*<0.001) were significantly higher in the DCI group compared with the non-DCI group. Both of those were also higher in the poor outcome group compared with the good outcome group, but the difference was not statistically significant at admission (0.53±0.18 vs0.49±0.14, *P*=0.198). K^trans^ in the non-DCI group (0.47±0.12 vs 0.41±0.14, *P*=0.004) and good outcome group (0.49±0.14 vs 0.41±0.14, *P*<0.001) decreased significantly from admission to DCITW. Multivariate analysis identified DCITW K^trans^ and admission K^trans^ as independent predictors of poor outcome (OR=1.73, 95%CI: 1.24-2.43, *P*=0.001) and DCI (OR=1.75, 95%CI: 1.25-2.44, *P*=0.001), respectively.

**Conclusion:** Elevated K^trans^ at admission is associated with the occurrence of DCI, but not with outcome at three months. Continuous monitoring of K^trans^ from admission to DCITW can accurately identify reversible and irreversible changes in K^trans^, and can predict outcome.

## Introduction

Aneurysmal subarachnoid hemorrhage (aSAH) accounts for 2-10% of all strokes worldwide. This pathology carries a high risk of early mortality and long-term disability in survivors.^1-4^ Early brain injury and delayed cerebral ischemia (DCI) are now recognized as major contributors to outcome following aSAH.^5^ Early brain injury occurs within 72 hours of ictus. The time window of DCI (DCITW) is defined as the time period between 4 to 21 days after aSAH.^4-6^ No clear causal link has been established between Early brain injury and DCI, however both factors determine long-term outcome.^5,7-10^ Therefore, early identification of Early brain injury severity and prediction of DCI represent critical aspects of successful aSAH management.^4^

It is well known that disruption of the blood-brain barrier (BBB) is a pathophysiological change common to ischemic and hemorrhage stroke.^11-13^ It occurs in the acute phase after the initial hemorrhage, and will last for a substantially longer period of time. This alteration is associated with brain edema, thrombosis, inflammation, and other pathophysiological events that may propel the development of DCI.^12^ Current guidelines^4,9^ have recommended the use of CT perfusion to access regions of potential brain ischemia after SAH. This methodology can also be used to evaluate BBB permeability (BBBP).^14^ Previous studies have focused on predicting DCI and its subsequent outcome via BBBP monitoring at different time points within three days of onset, resulting in inconsistent conclusions.^15-21^ Early BBB disruption in acute ischemic stroke is reversible, while persistent disruption is associated with poor outcome.^22, 23^ However, the exact time course of post-aSAH BBB disruption remains unclear. We hypothesize that BBBP may evolve in complex ways over time, thus playing different roles at different stages following the acute event.

In this study, we aimed to explore the prognostic value of BBBP at different stages using two strategies. First, we quantified BBBP within 24 hours of aSAH onset and during DCITW. We then compared differences in BBBP between DCI and non-DCI groups, and between good and poor outcome groups. We also evaluated changes in BBBP from the time of admission throughout DCITW in each group. Second, we explored the potential of BBBP at admission and during DCITW for predicting DCI and outcome prognosis at three months.

## Methods

This retrospective study was approved by the institutional review board of our institution. The need for informed consent was waived by the institutional review board due to the retrospective nature of this study. The data supporting the study findings are available upon reasonable request.

### Study Population

This retrospective analysis was conducted from our prospective database for aSAH. We recruited patients who were admitted to our institution between July 2020 and September 2022, and who met the following inclusion criteria: (1) age>18 years; (2) CT perfusion was performed within 24 hours of onset and during the DCITW; (3) aSAH was confirmed via non-contrast CT and digital subtraction angiography. Exclusion criteria were: (1) previous history of cerebrovascular disease;(2) poor image quality owing to motion artifacts or incomplete acquisition. All patients were treated in compliance with current guidelines^4,9^ and with the aSAH treatment protocol of our institution. Nimodipine was administered intravenously via a micropump during hospitalization.

All patient information was recorded at admission, including gender, age, previous history of hypertension, World Federation of Neurosurgery Scale (WFNS), Hunt-Hess grade (H-H), modified Fisher Score (mFS), and Subarachnoid Hemorrhage Early Brain Edema Score (SEBES).

### Analysis of 3-months outcome and DCI

The functional outcome at three months was scored using a modified Rankin scale, ranging between 0 and 6 points depending on the functional status of the patients.^24^ The mRS was recorded by a neurologist blinded to clinical and imaging data, during a clinical visit or telephone interview. Patients were classified into good (modified Rankin scale in the ranges 0-2) and poor (modified Rankin scale in the ranges 3-6) outcomes.

DCI was ascertained using the consensus definition^25^: (1) a focal neurological impairment or a decrease of at least two points on Glasgow Coma Scale, not apparent immediately after aneurysm occlusion, and not attributed to other conditions; (2) presence of cerebral infarction on CT or MRI, not present on imaging between 24 and 48 hours after aneurysm occlusion, and not attributed to other causes such as surgical clipping or endovascular treatment. The above clinical and imaging features were assessed by a radiologist and a neurologist, both blinded to the baseline CT perfusion result.

### Imaging Protocol and Post-processing

The whole-brain CT perfusion protocol (**Supplemental Figure S1**) was performed with a dual-source CT scanner (SOMATOM Definition Flash; Siemens Healthcare, Erlangen, Germany). A non-contrast CT scan (120 kV, 390 mAs, contiguous 5-mm axial sections) was first performed. Next, a 55-ml dose of non-ionic contrast agent (Ioversol, 350 mg I/ml) was intravenously injected through the cubital vein at a rate of 5ml/s using a double-tube high-pressure syringe (Ulrich, Buchbrunnenweg, Germany), followed by 40 ml of saline flush at the same rate. CT perfusion acquisition (80 kV, 100 mAs, collimation 32×1.2 mm, 150 mm along the z-axis) was started 5 s after the contrast agent was injected.

Post-processing was performed on the Siemens workstation (Siemens syngo.via VB10B) using CT neurovascular and CT Neuro Perfusion software. The shape of the arterial input function necessary for the Patlak analysis was automatically determined from branches of the middle cerebral artery or anterior cerebral artery. The peak of the input function was normalized to the peak of the superior sagittal sinus.^26^ Finally, the flow extraction product, also called K^trans^, was obtained from the Patlak algorithm, which models extravascular leakage of blood into the interstitial space to visualize BBB disturbances.^27^

### Imaging Analysis

After parameter calculation, thirty-two regions of interest were defined on five slices by taking into account artery supply (**Supplemental Figure S2**). The average value across regions of interest was used to characterize K^trans^, both at admission and during DCITW. The quantitative analysis was performed by a radiologist blinded to all other clinical and imaging data.

### Statistical Analysis

All statistical analysis were performed using SPSS version 25.0 (IBM, Armonk, NY, USA), and *P*<0.05 was adopted to indicate statistical significance. Quantitative variables are expressed as means ± standard deviation, or medians with interquartile ranges. We compared groups using independent *t* tests or Mann-Whitney *U* tests. We expressed categorical variables as frequencies (percentage). For these variables, we performed group comparisons using chi-square-test or Fisher’s exact probability tests. We compared K^trans^ between admission and DCITW using Paired *t* test. Variables with *P*<0.1 in the univariate analysis were included in the multivariate logistic regression analysis to identify independent predictors of DCI and poor outcome. We then incorporated the identified independent predictors for these two factors into their respective predictive models. Receiver operating characteristic curve analysis was performed to evaluate predictive performance.

## Results

### Patient characteristics

**Figure 1** shows a flowchart of patient selection. A total of 128 patients (mean age 61±12; 75 women) were included in this study. Aneurysms were repaired within 48 hours after symptom onset via coil embolization (94%) and microsurgical clipping (6%).

**Figure 1.**
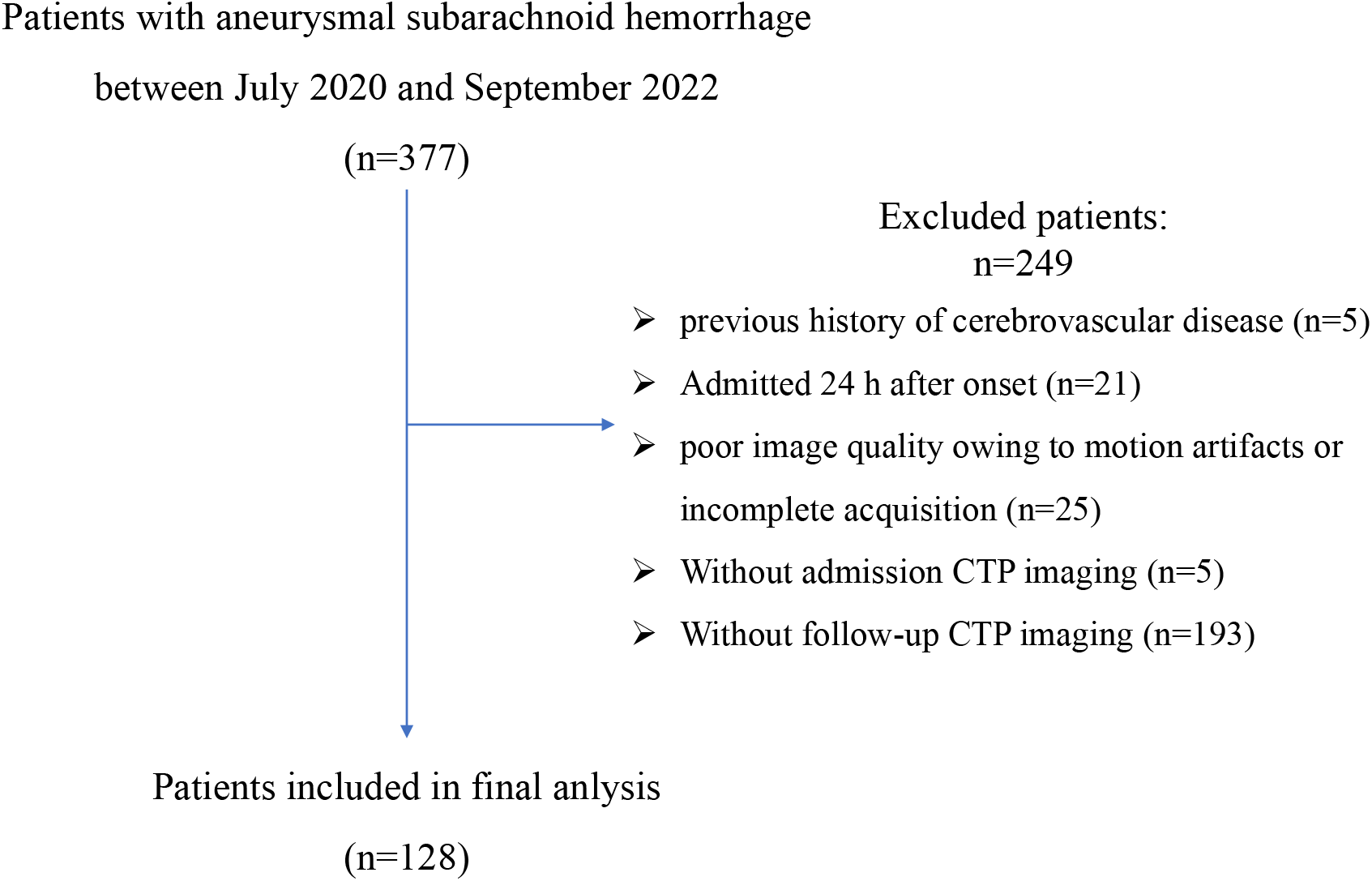
Flowchart of all patients.

### Comparison between non-DCI and DCI groups, and between good outcome and poor outcome groups, from admission to DCITW

**Table 1** summarizes relevant clinical and imaging information.

**Table 1:**
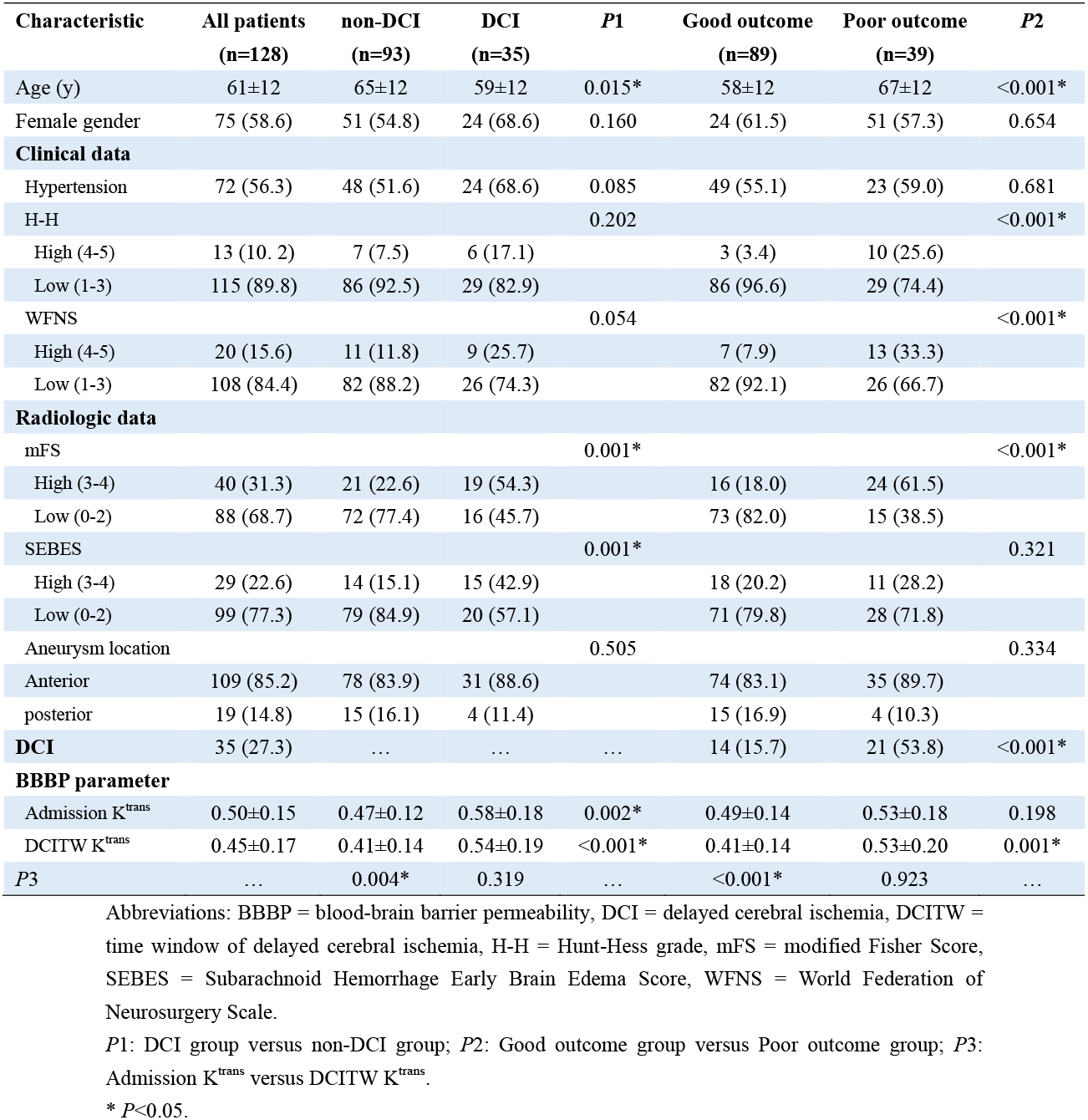
Comparison of demographic, clinical and imaging characteristics between non-DCI and DCI groups, and between good outcome and poor outcome groups, from admission to DCITW

| Characteristic | All patients<br>(n=128) | non-DCI<br>(n=93) | DCI<br>(n=35) | P1 | Good outcome<br>(n=89) | Poor outcome<br>(n=39) | P2 |
| --- | --- | --- | --- | --- | --- | --- | --- |
| Age (y) | 61±12 | 65±12 | 59±12 | 0.015* | 58±12 | 67±12 | <0.001* |
| Female gender | 75 (58.6) | 51 (54.8) | 24 (68.6) | 0.160 | 24 (61.5) | 51 (57.3) | 0.654 |
| **Clinical data** |  |  |  |  |  |  |  |
| Hypertension | 72 (56.3) | 48 (51.6) | 24 (68.6) | 0.085 | 49 (55.1) | 23 (59.0) | 0.681 |
| H-H |  |  |  | 0.202 |  |  | <0.001* |
| High (4-5) | 13 (10.2) | 7 (7.5) | 6 (17.1) |  | 3 (3.4) | 10 (25.6) |  |
| Low (1-3) | 115 (89.8) | 86 (92.5) | 29 (82.9) |  | 86 (96.6) | 29 (74.4) |  |
| WFNS |  |  |  | 0.054 |  |  | <0.001* |
| High (4-5) | 20 (15.6) | 11 (11.8) | 9 (25.7) |  | 7 (7.9) | 13 (33.3) |  |
| Low (1-3) | 108 (84.4) | 82 (88.2) | 26 (74.3) |  | 82 (92.1) | 26 (66.7) |  |
| **Radiologic data** |  |  |  |  |  |  |  |
| mFS |  |  |  | 0.001* |  |  | <0.001* |
| High (3-4) | 40 (31.3) | 21 (22.6) | 19 (54.3) |  | 16 (18.0) | 24 (61.5) |  |
| Low (0-2) | 88 (68.7) | 72 (77.4) | 16 (45.7) |  | 73 (82.0) | 15 (38.5) |  |
| SEBES |  |  |  | 0.001* |  |  | 0.321 |
| High (3-4) | 29 (22.6) | 14 (15.1) | 15 (42.9) |  | 18 (20.2) | 11 (28.2) |  |
| Low (0-2) | 99 (77.3) | 79 (84.9) | 20 (57.1) |  | 71 (79.8) | 28 (71.8) |  |
| Aneurysm location |  |  |  | 0.505 |  |  | 0.334 |
| Anterior | 109 (85.2) | 78 (83.9) | 31 (88.6) |  | 74 (83.1) | 35 (89.7) |  |
| posterior | 19 (14.8) | 15 (16.1) | 4 (11.4) |  | 15 (16.9) | 4 (10.3) |  |
| DCI | 35 (27.3) | ... | ... | ... | 14 (15.7) | 21 (53.8) | <0.001* |
| **BBBP parameter** |  |  |  |  |  |  |  |
| Admission $K^{trans}$ | 0.50±0.15 | 0.47±0.12 | 0.58±0.18 | 0.002* | 0.49±0.14 | 0.53±0.18 | 0.198 |
| DCITW $K^{trans}$ | 0.45±0.17 | 0.41±0.14 | 0.54±0.19 | <0.001* | 0.41±0.14 | 0.53±0.20 | 0.001* |
| P3 | ... | 0.004* | 0.319 | ... | <0.001* | 0.923 | ... |

Abbreviations: BBBP = blood-brain barrier permeability, DCI = delayed cerebral ischemia, DCITW = time window of delayed cerebral ischemia, H-H = Hunt-Hess grade, mFS = modified Fisher Score, SEBES = Subarachnoid Hemorrhage Early Brain Edema Score, WFNS = World Federation of Neurosurgery Scale.
P1: DCI group versus non-DCI group; P2: Good outcome group versus Poor outcome group; P3: Admission $K^{trans}$ versus DCITW $K^{trans}$ .
\* $P < 0.05$ .

Twenty-seven percent (35/128) of patients developed DCI during hospitalization. Patients with DCI were older (*P*=0.015) and presented higher SEBES (*P*=0.001) and mFS (*P*=0.001) than those without DCI at admission. Both admission K^trans^ (*P*=0.002) and DCITW K^trans^ (*P*<0.001) in the DCI group were significantly higher than those in the non-DCI group. K^trans^ in the non-DCI group decreased significantly from the time of admission throughout DCITW (0.47±0.12 versus 0.41±0.14, *P*=0.004; **Figures 2A**), but not in the DCI group (0.58±0.18 versus 0.54±0.19, *P*=0.319; **Figures 2B**).

**Figure 2.**
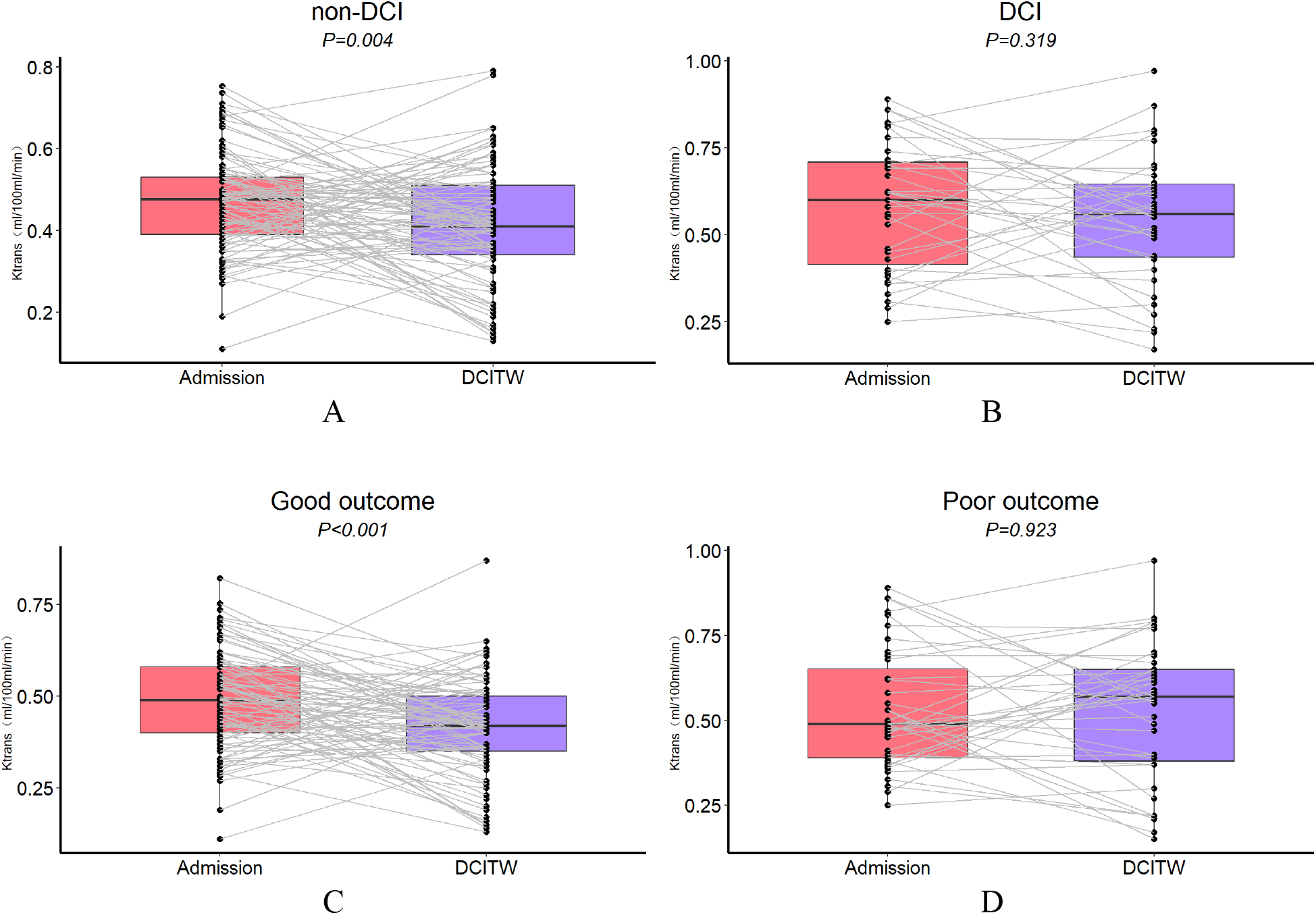
Diagrams show changes in K^trans^ from the time of admission throughout delayed cerebral ischemia (DCI) time window (DCITW).K^trans^ in the non-DCI group (0.47±0.12 versus 0.41±0.14, *P*=0.004; A) and good outcome group (0.49±0.14 versus 0.41±0.14, *P*<0.001; C) decreased significantly from the time of admission throughout DCITW, but not in the DCI group (0.58±0.18 versus 0.54±0.19, *P*=0.319; B) and poor outcome group (0.53±0.18 versus 0.53±0.20; *P*=0.923; D).

Thirty percent (39/128) of patients had poor outcome at three months. Patients with poor outcome were older and presented higher WFNS, H-H, and mFS values than those with good outcome at admission (all *P*<0.001). Both admission K^trans^ (*P*=0.198) and DCITW K^trans^ (*P*=0.001) in the poor outcome group were higher than corresponding values in the good outcome group, but there was no statistical difference at admission. K^trans^ in the good outcome group decreased significantly from the time of admission throughout DCITW (0.49±0.14 versus 0.41±0.14, *P*<0.001; **Figures 2C**), but not in the poor outcome group (0.53±0.18 versus 0.53±0.20; *P*=0.923; **Figures 2D**).

Notably, the majority (60%) of patients with DCI presented poor outcome at three months (*P*<0.001; **Figure 3**).

**Figure 3.**
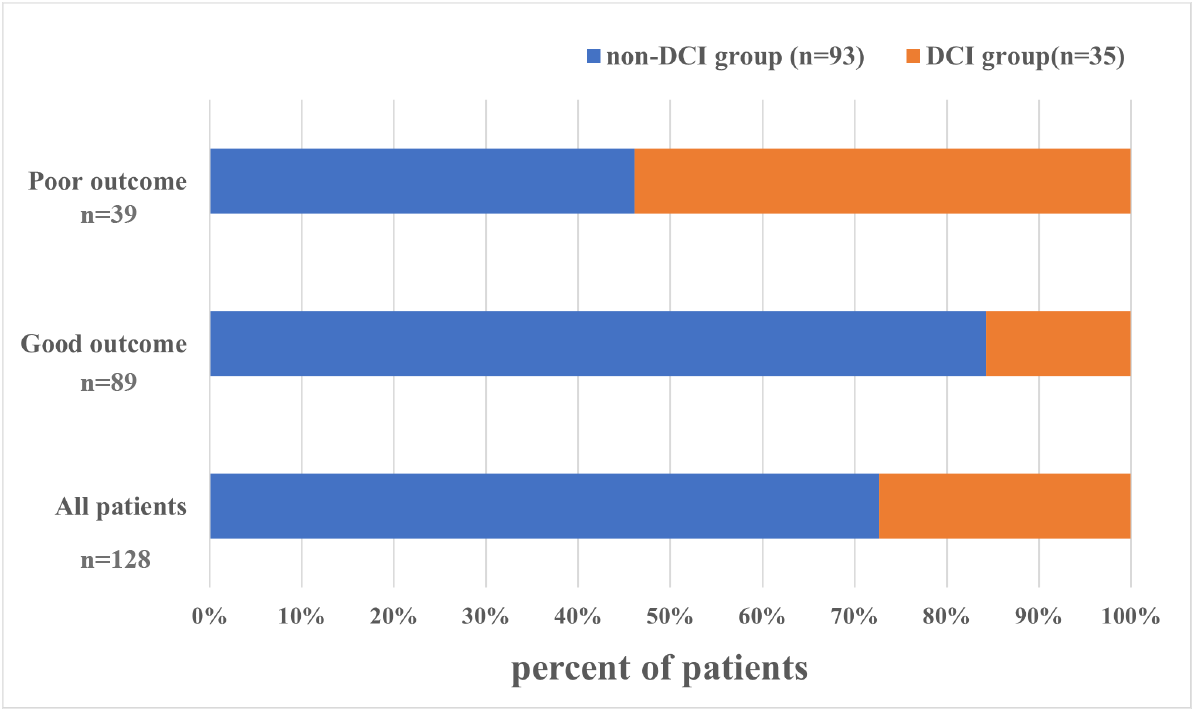
Distribution of delayed cerebral ischemia (DCI) among patients in the good outcome group and poor outcome group.The majority (60%) of patients with DCI presented poor outcome at three months.

### Predictive performance of clinical and imaging data

Multivariable logistic regression analysis showed that admission K^trans^ (OR=1.75, 95%CI: 1.25-2.44, *P*=0.001), age (OR=1.07, 95%CI: 1.02-1.12, *P*=0.002), SEBES (OR=5.63, 95%CI: 1.87-16.93, *P*=0.002), and mFS (OR=2.71, 95%CI: 1.04-7.10, *P*=0.042) were independent predictors of DCI. Receiver operating characteristic curve analysis revealed that the predictive model for DCI produced an AUC of 0.832 (95%CI: 0.75-0.91; **Figures 4A**).

**Figure 4.**
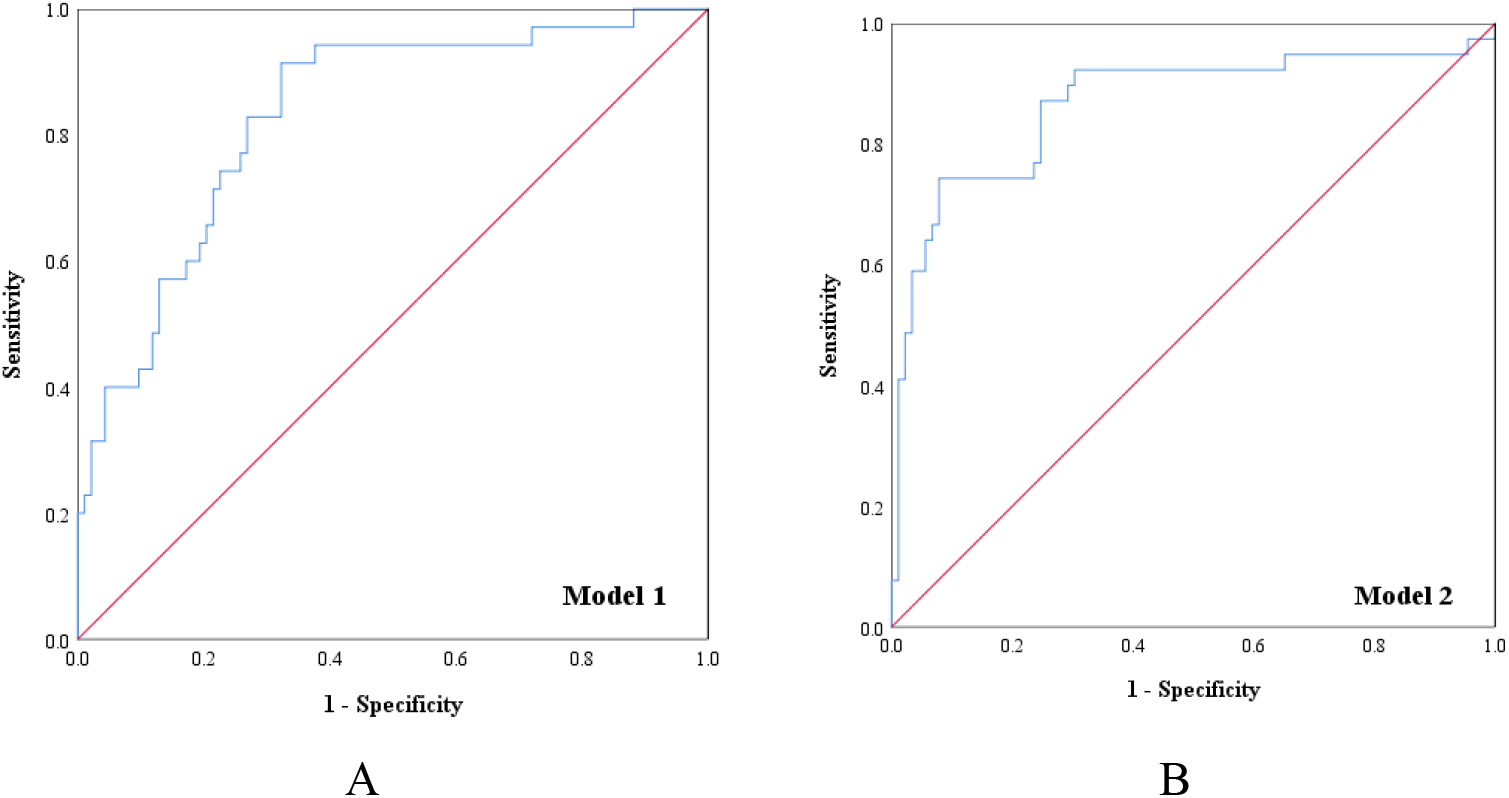
Receiver operating characteristic curves show sensitivity and specificity of Model 1 and Model 2 for prediction of delayed cerebral ischemia (A) and poor outcome (B), respectively.

Moreover, DCITW K^trans^ (OR=1.73, 95%CI: 1.24-2.43, *P*=0.001), age (OR=1.06, 95%CI: 1.01-1.11, *P*=0.013), mFS (OR=5.72, 95%CI: 1.99-16.42, *P*=0.001), and H-H (OR=7.12, 95%CI: 1.17-43.19, *P*=0.033) were independent predictors of poor outcome. The predictive model for poor outcome produced an AUC of 0.867 (95%CI: 0.79-0.95; **Figures 4B**).

## Discussion

In the present study, we investigated the prognostic value of BBBP at admission and during DCITW following aSAH. We found the following results. First, elevated K^trans^ at admission is associated with the occurrence of DCI, but not with outcome at three months. Second, K^trans^ decreased significantly in the non-DCI group and good outcome group from the time of admission and throughout DCITW, but this trend was not present for the DCI group and poor outcome group. Third, continuously elevated K^trans^ during the DCITW is a predictor of poor outcome.

Previous studies have evaluated how BBBP may be connected with subsequent DCI and long-term outcome in aSAH patients.^15-21^ However, the resulting conclusions remain controversial with regard to the value of using BBBP to predict DCI and outcome. This controversy may reflect inconsistencies in the time point of imaging acquisition, as previous studies have not assessed the time evolution of BBBP. Animal models have demonstrated that BBB disruption evolves over time, beginning with ischemia and worsening with sustained hypoperfusion.^28^ In line with these results, we demonstrated here that BBBP changes its characteristics at different stages following aSAH, thus carrying different prognostic value depending on time of acquisition. K^trans^ at the time of admission may reflect the extent of early brain injury, which is closely associated with DCI. Consistently elevated values of K^trans^ during DCITW may instead reflect the development of DCI, and serve as predictors of poor outcome. An additional source of potential inconsistency across previous studies is represented by their relatively small sample sizes, ranging between 16 and 69. In some studies, patients presented relatively good clinical conditions at admission. Both factors may influence the correlation between BBBP and clinical outcome.^21^

Prior study with a small sample size showed that early moderate BBB disruption may be reversible, thus limiting sustained brain injury, whereas severe BBB disruption may be irreversible, and therefore associated with DCI.^15^ The present results further support this hypothesis: we found that changes in K^trans^ from the time of admission throughout DCITW differed between DCI and non-DCI groups, and between good and poor outcome groups. K^trans^ decreased significantly in the non-DCI group and in the good outcome group, but not in the DCI and poor outcome groups. This result points to the existence of both reversible and irreversible changes in K^trans^ after aSAH. At the same time, the majority (60%) of patients with DCI presented poor outcome at three months. This finding suggests that irreversible changes occur more often in patients with DCI, resulting in a poor outcome. This interpretation also helps explain why multivariate analysis showed that admission K^trans^ is an independent predictor of DCI, while DCITW K^trans^ is an independent predictor of poor outcome. Recently, a randomized controlled trail demonstrated that the use of goal-directed hemodynamic therapy significantly reduces the occurrence of DCI, and ensures better prognosis.^29^ These results further emphasize the importance of identifying reversible and irreversible changes in BBBP for predicting DCI and functional outcome.

In addition to clarifying the role of admission and DCITW K^trans^, our findings are also in line with previous studies suggesting that age, SEBES, and mFS are independent predictors of DCI,^30,31^ while age, H-H, and mFS are independent predictors of poor outcome.^11,32^ All the predictors mentioned above can be quickly obtained in a short period of time, allowing for a simple and feasible prediction model when combined with K^trans^ at admission or during DCITW.

Our study presents several limitations. First, the relatively small sample size and retrospective design may have introduced selection bias, and make it difficult to conduct sub-group analysis based on neurological grade. Second, CT perfusion was performed only at admission and during DCITW. A protocol with standardized time points for imaging acquisition may be more accurate in assessing BBBP changes and their prognostic significance.

## Conclusions

Our results indicate that the characteristics of BBBP evolve over different stages following aneurysmal subarachnoid hemorrhage, thus carrying different prognostic value over time. Elevated K^trans^ at admission is associated with the occurrence of DCI, but not with outcome at three months. Continuous assessment of K^trans^ from admission to DCITW can accurately identify a reversible or irreversible change in BBBP, and can predict outcome at three months. It is therefore important to characterize BBBP at different stages after the initial acute event, and to distinguish whether it reflects reversible or irreversible changes that may inform effective planning of subsequent clinical care.

## Data Availability

The data supporting the study findings are available upon reasonable request.

None

None

## Non-Standard Abbreviations and Acronyms

aSAH: aneurysmal subarachnoid hemorrhage,
BBB: blood-brain barrier,
BBBP: blood-brain barrier permeability,
DCI: delayed cerebral ischemia,
DCITW: time window of delayed cerebral ischemia,
H-H: Hunt-Hess grade,
mFS: modified Fisher Score,
SEBES: Subarachnoid Hemorrhage Early Brain Edema Score,
WFNS: World Federation of Neurosurgery Scale

## Acknowledgement

We thank Peter Neri, from Liwen Bianji (Edanz) (www.liwenbianji.cn) for editing the English text of a draft of this manuscript.

## Supplemental material

Supplemental figures 1 and 2.

